# Outcomes of COVID-19 infection in patients treated with Clozapine

**DOI:** 10.1101/2021.04.13.21255384

**Authors:** Risha Govind, Daniela Fonseca de Freitas, Megan Pritchard, Mizanur Khondoker, James T Teo, Robert Stewart, Richard D. Hayes, James H. MacCabe

## Abstract

**Background:** Clozapine, an antipsychotic, is associated with increased susceptibility to infection with COVID-19, compared to other antipsychotics.

**Aims:** To investigate associations between clozapine treatment and increased risk of adverse outcomes of COVID-19, namely COVID-related hospitalisation and intensive care treatment, and death, among patients taking antipsychotics with schizophrenia-spectrum disorders.

**Method:** Using data from South London and Maudsley NHS Foundation Trust (SLAM) clinical records, via the Clinical Record Interactive Search system, we identified 157 individuals who had an ICD-10 diagnosis of schizophrenia-spectrum disorders, were taking antipsychotics at the time of the COVID-19 pandemic in the UK, and had a laboratory-confirmed COVID-19 infection. The following health outcomes were measured: COVID-related hospitalisation, COVID-related intensive care treatment death. We tested associations between clozapine treatment and each outcome using logistic regression models, adjusting for gender, age, ethnicity, neighbourhood deprivation, obesity, smoking status, diabetes, asthma, bronchitis and hypertension using propensity scores.

**Results:** In the 157 individuals who developed COVID while on antipsychotics, there were 44 COVID-related hospitalisations, 13 COVID-related intensive care treatments and 13 deaths of any cause during the follow-up period. In the unadjusted analysis, there was no significant association between clozapine and any of the outcomes and there remained no associations following adjusting for the confounding variables.

**Conclusions:** In our sample of patients with COVID-19 and schizophrenia-spectrum disorders, we found no evidence that clozapine treatment puts patients at increased risk of hospitalisation, intensive care treatment or death, compared to any other antipsychotic treatment. However, further research should be considered in larger samples to confirm this.

**Conflict of interest:** RDH has received research funding from Roche, Pfizer, Janssen, and Lundbeck. DFF has received research funding from Janssen and Lundbeck. JHM has received research funding from Lundbeck. JTT has received research funding from Bristol-Meyers-Squibb. RS declares research support in the last 36 months from Janssen, GSK and Takeda.

**Ethics statement:** The research was conducted under ethical approval reference 18/SC/0372 from Oxfordshire Research Ethics Committee C.

## INTRODUCTION

Clozapine is an atypical antipsychotic, the gold standard drug for treatment-resistant schizophrenia, and the only effective treatment for many patients with schizophrenia (1). Patients with schizophrenia have a higher risk for developing pneumonia and, compared to the general population, have higher premature mortality (2–7). Patients receiving clozapine treatment have lower rates of overall hospitalisation and mortality compared to those receiving other antipsychotic treatments (8–11). However, clozapine is associated with an increased risk of developing pneumonia (12–15). This might be explained by confounding by indication, in that clozapine is predominantly prescribed in cases of treatment-resistant schizophrenia, associated in itself with higher rates of comorbidities such as smoking cigarettes, inadequate physical activity, and poor diet (16). Alternatively, clozapine could increase the risk of pneumonia via immunosuppression, or via other adverse effects of clozapine which could fall on the causal pathway, such as hypersalivation (causing aspiration pneumonia), diabetes and obesity (15–17). COVID-19 first appeared in China in December 2019 and was declared a global pandemic by the WHO in March 2020 (18). It is caused by the SARS-Cov2 virus, and has pathological effects on multiple organ systems including the lungs, heart, brain, kidney, gastrointestinal tract, liver and spleen (19). The most concerning consequence of the infection is respiratory failure. The most severe cases of COVID-19 can require hospitalisation and treatment in intensive care, and mortality is significant. In a previous study we reported that patients on clozapine treatment may be at higher risk of COVID-19 infection (20). Recently, case studies on this have been presented by Butler et al., Boland and Dratcu (21,22); however, to our knowledge, the association between clozapine treatment and the adverse outcomes of COVID-19 have yet to be investigated in an epidemiological sample. In this paper, we investigated whether clozapine treatment was associated with an increased risk of adverse outcomes of COVID-19 in patients with schizophrenia and other psychoses treated with antipsychotics in a geographically defined population in London during the COVID-19 pandemic in the UK.

## METHOD

### Setting and Ethics Statement

A retrospective cohort study was carried out using data from the electronic records of the South London and Maudsley NHS Foundation Trust (SLAM). SLAM caters to all secondary mental health care needs of over 1.3 million people of four London boroughs (Lambeth, Southwark, Lewisham, and Croydon). SLAM has used a fully electronic clinical records system since 2006, and the Clinical Records Interactive Search (CRIS) platform was established to render full, de-identified clinical records available to researchers for secondary analysis within a robust data security and governance framework (23). CRIS was approved for use as a de-identified data resource for secondary analysis by Oxfordshire Research Ethics Committee C (reference 18/SC/0372).

CRIS includes both structured and free-text fields from the clinical notes, and custom-built Natural Language Processing (NLP) algorithms are used to extract information from the latter, the specifications and performance metrics of which are detailed in an open online catalogue (24). Data from four NLP algorithms were used in this study: diagnosis, medication, smoking and body mass index (BMI). Information regarding COVID-19 patient cases admitted to two local hospitals (King’s College Hospital and Princess Royal University Hospital) were obtained via a data linkage (performed under Regulation 3(2) and Regulation 3(3) of the Health Service Control of Patient Information Regulations 2002 (COPI)).

### Cohort

The cohort comprised individuals who satisfied all three of the following inclusion criteria: (1) a laboratory-confirmed COVID-19 infection between March 01, 2020 and December 20, 2020; (2) ICD-10 diagnosis of any schizophrenia-spectrum disorder (F2*); (3) recorded as taking antipsychotic medication within 3 months prior to the date of COVID-19 infection. Figure 1 shows the study design. SQL Server Management Studio version 15.0 (Microsoft Inc, USA) was used to extract the data. The day of data extraction was January 07, 2021. Patients were followed-up from the date of COVID-19 infection until they were hospitalised, entered intensive care treatment, died, or reached the end of the follow-up period (within 28 days of infection).

**Figure 1:**
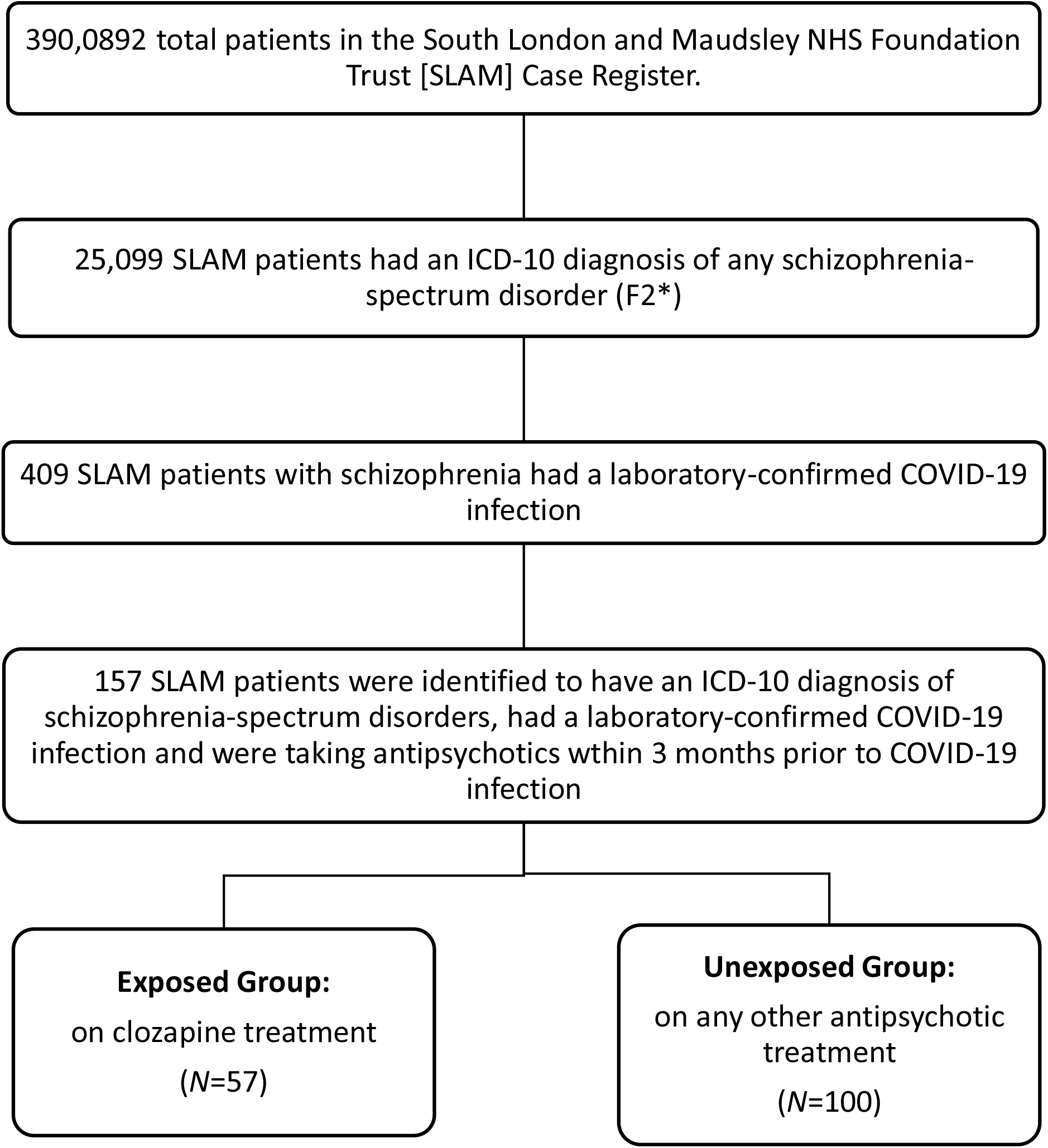
Study design

Diagnosis of schizophrenia-spectrum disorder (ICD-10: F2*) was ascertained via a diagnosis algorithm, by which NLP outputs are combined with data in the structured fields, such as the data from ICD-10 diagnosis forms in the source record (24).

Antipsychotic medication within 3 months prior to COVID-19 infection was identified by an NLP algorithm which targeted administrations of clozapine, amisulpride, aripiprazole, chlorpromazine, droperidol, flupentixol, fluphenazine, haloperidol, lurasidone, olanzapine, paliperidone, piportil, pipotiazine, quetiapine, risperidone, sulpiride, trifluoperazine and zuclopenthixol (24). The medications algorithm NLP outputs are combined with data from structured fields, including SLAM pharmacy dispensing data.

The COVID-19 infection data used for the inclusion criteria were collated by combining information from three sources: (1) SLAM pathology lab results data, (2) the presence of a clinician-entered alert on SLAM records indicating a positive test, and (3) data provided by local general hospitals (King’s College Hospital and Princess Royal University Hospital) for COVID-19 related admissions. The COVID-19 infection dates were verified and, when needed, were rectified to the earliest mention of COVID-19-compatible symptoms or COVID-19 tests, according to the information presented in SLAM’s clinical notes. To cater to scenarios where the COVID-19 test was conducted after an admission for symptomatic COVID, the COVID-19 infection date was changed to the date of hospital admission when the positive test result was within 7 days of hospital admission. The patients were removed from the analysis either if the clinical notes stated that their COVID-19 positive status was entered by mistake or they had COVID-19 infection after December 20, 2020.

### Exposure of interest

People who were recorded as receiving clozapine treatment at any time within 3 months prior to the assigned COVID-19 infection date were defined as the exposed group. Those on any type or combination of antipsychotic treatment that did not include clozapine during this time constituted the unexposed group.

### Main outcome measures

The outcomes of interest were: (1) COVID-related hospitalisation (2) COVID-related intensive care treatment, and (3) all-cause mortality during the follow-up period (within 28 days of COVID-19 infection). These data were collated by combining COVID-19 related information provided by local general hospitals (King’s College Hospital and Princess Royal University Hospital) and the data in the SLAM records. The SLAM records data on hospitalisation and intensive care treatment were curated by reading the clinical notes of each patient from the date of COVID-19 infection until a positive mention of hospitalisation or mention of recovery from COVID-19. The SLAM records data on mortality were retrieved from structured fields in SLAM health records which are populated on weekly basis via linkage with the NHS Spine.

### Potential confounding variables

We considered as potential confounding variables sociodemographic characteristics and behavioural/clinical factors. The sociodemographic information comprised age, gender, ethnicity, and neighbourhood deprivation. The behavioural/clinical factors were smoking status, obesity, diabetes, asthma, bronchitis and hypertension.

Age was calculated at the time of COVID-19 infection from the year and month of birth. Data on gender and ethnicity came from the routinely collected data in structured fields in SLAM health records. SLAM records include ethnicity in 14 categories, which were collapsed into 3 categories, “White”, “Black” and “Asian & other”. The category “White” was a conflation of White British, White Irish and White Other. The category “Black” was a conflation of Black African, Black Other (which comprises Black British), Black Caribbean, Mixed Race White and Black Caribbean and Mixed Race White and Black African. The category “Asian & Other” was a conflation of Indian, Pakistani, Other Asian, and Other ethnic group. For patients with no ethnicity information, including those with ethnicity as “not stated” in the structured fields, their ethnicity data was extracted by manually reviewing the record text fields.

Neighbourhood deprivation was measured using the Index of Multiple Deprivation (IMD) 2019 applying Census-derived data to the Lower Super Output Area: a standard administrative unit containing an average of 1500 residents. The deciles of the IMD range between 1, the most deprived, and 10, the least deprived. The data from IMD deciles 1 to 3 were merged to form the “Higher level of deprivation” category. The data from IMD deciles 4 to 10 were merged to form the “Lower level of deprivation” category. A third category, “homeless”, was created for the patients who were homeless.

Smoking behaviour in the year prior to COVID-19 infection was identified using an NLP algorithm (24), supplemented by manual review of record text fields. Similarly, the obesity status (BMI≥30) was derived from recorded BMI scores ascertained via an NLP algorithm, supplemented by manual records text review, choosing the most recent extracted score prior to the COVID-19 infection date (24). Data on physical illnesses (diabetes, asthma, bronchitis and hypertension) were extracted manually from relevant free-text fields of the patient records for each patient, aided by search strings.

### Statistical analysis

The data were analysed using STATA for Windows version 15.1. Since the data on the date of COVID-19 infection which is the time zero date was not precise, we used logistic regression instead of Cox proportional hazard models for the analysis. In the unadjusted analysis, we used logistic regression to calculate odds ratios comparing clozapine treated patients to those treated with other antipsychotics for each of the outcomes described above. Covariate adjustment was made via propensity scores within a logistic regression model as direct adjustment for all covariates was not feasible due to limited sample size. The propensity scores were predicted from a separate logistic regression model using clozapine treatment as the outcome and the sociodemographic (age, gender, ethnicity, neighbourhood deprivation), behavioural/clinical factors (smoking status, BMI, diabetes, asthma, bronchitis, hypertension) as predictor variables. The logit (log-odds) of the probability of clozapine treatment (propensity score) was included as a single covariate along with the exposure (indicator of clozapine treatment) in the logistic regression models for adjusted analysis. STATA was also used to estimate power for the analysis.

## RESULTS

There were 157 patients ascertained with a laboratory-confirmed COVID-19 infection and schizophrenia-spectrum disorders (F2*) who were receiving any type of antipsychotic treatment during the study period. The follow-up period was 28 days after COVID-19 infection. The mean age of the study sample was 50.6 years (SD=16.01), and men accounted for 53.5% of the sample. The study sample had a relatively high proportion of patients from minority ethnic groups: 61.2% Black, 10.8% any Asian and Other ethnic background and 28.0% White. Table 1 summarises the demographic features of all the SLAM patients who were eligible for inclusion based on the inclusion criteria (*N*=157). Of the individuals who were receiving clozapine, 56% were male, 58% were Black, 88% were current smokers, and 63% were obese.

**Table 1:** Sample description of the 157 SLAM patients who qualified for the inclusion criteria, presented according to those who were and were not receiving clozapine-treatment

|  | On Clozapine<br>treatment<br>(%) | Not on Clozapine-treatment<br>(%) |
| --- | --- | --- |
| Total Patients in cohort | 36.3 (n=57) | 63.7 (n=100) |
| Males | 56.1 | 52.0.0 |
| Age |  |  |
| < 50 years | 52.6 | 40.0 |
| > 50 years | 47.4 | 60.0 |
| Ethnicity |  |  |
| White | 28.1 | 28.0 |
| Black | 57.9 | 63.0 |
| Asian & Other | 14.0 | 9.0 |
| Neighbourhood Deprivation |  |  |
| Lower level of deprivation | 56.1 | 45.0 |
| Higher level of deprivation | 35.1 | 49.0 |
| Homeless | 8.8 | 6.0 |
| Current smoker | 87.7 | 58.0 |
| Obesity | 63.2 | 35.0 |
| Diabetes | 43.9 | 42.0 |
| Asthma | 28.1 | 16.0 |
| Bronchitis | 8.8 | 13.0 |
| Hypertension | 45.6 | 45.0 |

Of the 157 individuals, 44 had an episode of COVID-related hospitalisation and 13 received COVID-related intensive care treatment during the follow-up period. The patients receiving intensive care treatment were a subset of the 44 patients with COVID-related hospitalisation. Of the 13 patients receiving intensive care treatment, 3 died. All outcomes occurred within the follow-up period of 28 days since infection.

The logistic regression analysis was performed on the 157 individuals for each of the three outcomes, and Table 2 shows the odds ratio for each in the unadjusted and propensity score adjusted models. In unadjusted analyses, receiving clozapine-treatment was not significantly associated with any outcome. Furthermore, no significant association was observed for any of the outcomes after covariate adjustment. Post-hoc power calculations indicated that the sample size was sufficient to detect with 80% power (alpha 0.05) an odds ratio of 2.78 for COVID-related hospitalisation and 3.95 for all-cause mortality.

**Table 2:**
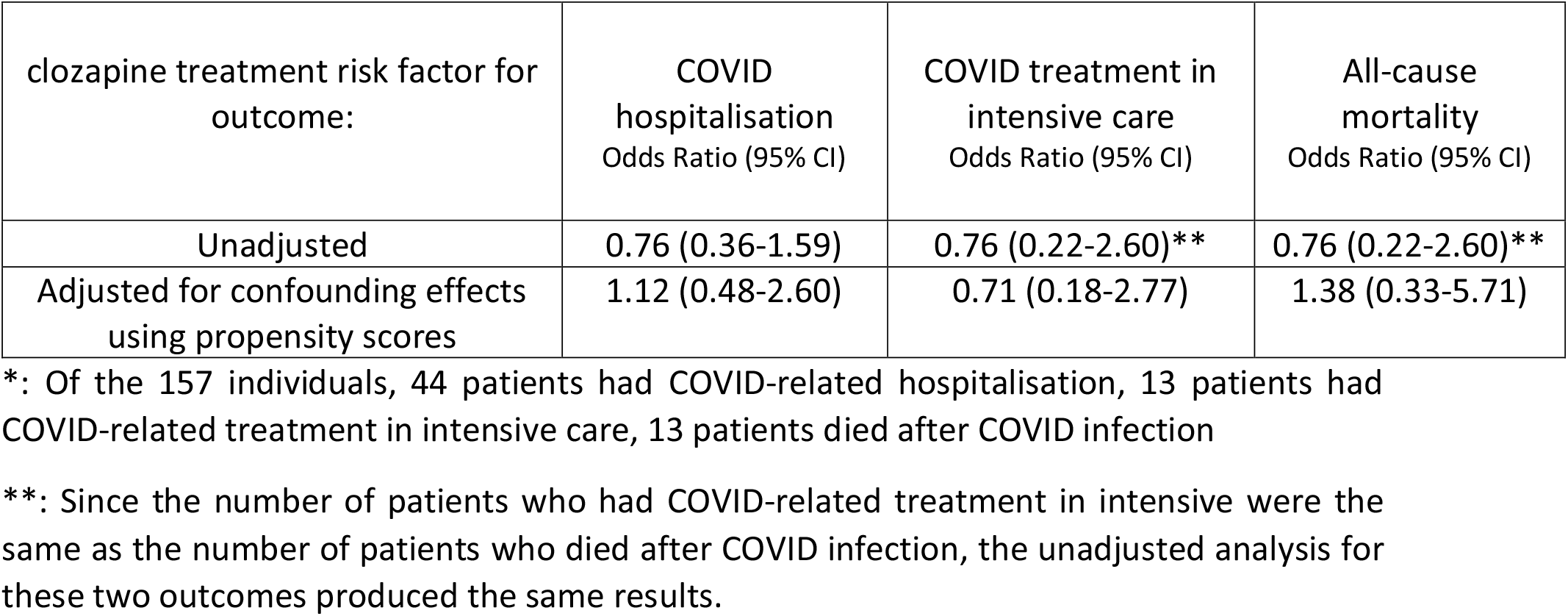
Logistic regression analysis of association between receiving clozapine treatment and each outcome (COVID-related hospitalisation, COVID-related treatment in intensive care and death) between the date of COVID infection and January 07, 2021, inclusive in 157 individuals*

| clozapine treatment risk factor for outcome: | COVID hospitalisation<br>Odds Ratio (95% CI) | COVID treatment in intensive care<br>Odds Ratio (95% CI) | All-cause mortality<br>Odds Ratio (95% CI) |
| --- | --- | --- | --- |
| Unadjusted | 0.76 (0.36-1.59) | 0.76 (0.22-2.60)** | 0.76 (0.22-2.60)** |
| Adjusted for confounding effects using propensity scores | 1.12 (0.48-2.60) | 0.71 (0.18-2.77) | 1.38 (0.33-5.71) |

\*: Of the 157 individuals, 44 patients had COVID-related hospitalisation, 13 patients had COVID-related treatment in intensive care, 13 patients died after COVID infection
\*\* : Since the number of patients who had COVID-related treatment in intensive were the same as the number of patients who died after COVID infection, the unadjusted analysis for these two outcomes produced the same results.

## DISCUSSION

### Summary of findings

We investigated if receiving clozapine treatment may be associated with increased risk of hospitalisation, intensive care treatment or all-cause mortality (within 28 days from infection) in COVID-19 positive patients with schizophrenia-spectrum disorders. We found no evidence that receiving clozapine treatment substantially increases the risk of these outcomes, compared to receiving any other types of antipsychotic treatment.

### Comparison with previous studies

To our knowledge, no previous research has specifically investigated the associations between receiving clozapine treatment, as compared to receiving treatment with other antipsychotics, and hospitalisation, intensive care treatment or mortality from COVID-19.

### Strengths and limitations

As a strength of this study, SLAM is a near-monopoly service provider of all aspects of secondary mental health care to residents within a defined geographic catchment, allowing relatively comprehensive ascertainment of people with the disorders of interest receiving specialist care during the COVID-19 pandemic in the UK. The CRIS database provided the platform to ascertain the relevant sample and access information on a range of potential confounders. However, it is important to bear in mind that not all people with schizophrenia-spectrum disorders will have been receiving specialist mental healthcare at that time, so that generalisability is limited. Furthermore, not all COVID-19 infection episodes will have been ascertained, particularly during early stages of the pandemic when access to tests was very limited. Another important limitation of the analysis results from type II error. We acknowledge this analysis is underpowered, although the urgency of the situation led us to decide to present data available at this stage.

The COVID-related hospitalisation and intensive care treatment data came from combining information provided by local general hospitals and supplementing that by reading clinical notes of each patients. For patients who did not receive COVID-related treatments at the local general hospitals, we have assumed their clinical notes would capture mentions of any COVID-related treatments they received. However, it cannot be ruled out that this might have been missed is some instances. Although all deaths occurred within 28 days of COVID infection, and thus meet the Public Heath England criteria for COVID-related deaths (25), it is possible that some deaths may have been unrelated to COVID.

Obesity is a recognised risk factor for more severe COVID-19 outcomes, but obesity information had to be extrapolated using the nearest BMI score, not all of which were recent. While these BMI scores are likely to give some indication of obesity of the patient, we cannot rule out that clozapine-treated patients may have more up-to-date BMI scores due to increased monitoring.

Since the smoking data encompasses patients who have mentions of cigarette smoking in their clinical records from any time within a year prior to COVID infection, some data were from almost a year ago. Given the impact of smoking on clozapine metabolization and clozapine plasma levels, it is important to note that clozapine-treated patients are more likely to be questioned about their smoking habits and therefore have recent information on it.

### Implications

To our knowledge, this is the first study to investigate whether patient’s clozapine-treated patients are at increased risk of adverse outcomes of COVID-19, such as hospitalisation, treatment in intensive care or ventilation, or all-cause mortality. Within the limits of statistical power, we did not find evidence of substantial increased risk; however, larger and/or multi-site studies would be needed to rule out smaller effects.

## Data Availability

This is a retrospective cohort study was carried out using data from the electronic records of the South London and Maudsley NHS Foundation Trust (SLAM). This data cannot be made available. The analysis used a de-identified data resource approved by Oxfordshire Research Ethics Committee C (reference 18/SC/0372).

